# Diabetic retinopathy screening practice and associated factors among adults with diabetes mellitus at a tertiary hospital in Addis Ababa, Ethiopia: a cross-sectional study

**DOI:** 10.64898/2026.11.10.26367772

**Authors:** Bezawit Shawel Moreda, Solomon Getachew Alem, Walelegn Yallew Worku

## Abstract

Diabetic retinopathy is a leading cause of preventable blindness. Because it is asymptomatic until substantial damage has occurred, periodic retinal examination is essential for early detection. Evidence on uptake where eye services are co-located with diabetes care in Ethiopia is lacking. This study assessed the prevalence of, and factors associated with, adequate diabetic retinopathy screening practice among adults with diabetes attending Menelik II Comprehensive Specialized Hospital, Addis Ababa.

We conducted an institution-based cross-sectional study from December 2023 to February 2024 among adults with type 1 or type 2 diabetes on follow-up for at least three months, selected by systematic random sampling and interviewed with a structured questionnaire. Knowledge and attitude were classified using a modified Bloom’s cut-off of 80%, and screening practice as adequate when at least two of three recommended practices were met. Factors associated with adequate practice were identified using multivariable logistic regression.

Of 404 patients approached, 371 were interviewed and included in the analysis (response rate 91.8%). Adequate screening practice was found in 139 participants (37.5%, 95% confidence interval [CI] 32.7–42.5); 62.0% had adequate knowledge and 72.8% a favorable attitude. Adequate practice was associated with adequate knowledge (adjusted odds ratio [AOR] 5.10, 95% CI 2.81– 9.25), a known eye disorder (AOR 3.96, 95% CI 2.11–7.43), higher education (AOR 3.82, 95% CI 1.33–10.95), age 41–65 years (AOR 3.15, 95% CI 1.10–9.03) and being married (AOR 1.73, 95% CI 1.01–2.97). Cost, lack of time and travel distance were the most commonly reported barriers.

Fewer than four in ten adults with diabetes underwent retinal examination at the recommended frequency despite on-site specialist eye care. The strong association with existing eye disorders suggests that examination is often prompted by ocular problems rather than undertaken for prevention. Strengthening patient education during diabetes follow-up and a direct referral pathway to on-site eye services may improve uptake.

## Introduction

Diabetes mellitus is a group of chronic metabolic disorders in which persistent hyperglycemia, arising from deficient insulin secretion, impaired insulin action, or both, damages the microvasculature and macrovasculature of multiple organ systems [1]. In the eye, this damage manifests as diabetic retinopathy, a progressive condition that remains asymptomatic until the retina is already substantially damaged and that is among the leading causes of preventable blindness in working-age adults [2]. Globally, an estimated 103 million people were living with diabetic retinopathy in 2020, a figure projected to reach 161 million by 2045, with the largest increases expected in low- and middle-income countries [3].

Ethiopia is experiencing this transition. A national systematic review and meta-analysis estimated the pooled prevalence of diabetic retinopathy among people with diabetes at 19.5% [4], while facility-based studies have reported estimates ranging from 13% in southern Ethiopia [5] to 34% in the north-west of the country [6]. Because early retinopathy produces no symptoms, it can only be identified through periodic examination of the retina. International and regional guidance therefore recommends that every person with type 1 or type 2 diabetes undergo a dilated retinal examination at least annually, with more frequent review once retinopathy is present [7,8]. Timely detection allows laser photocoagulation or intra-vitreal therapy to be delivered while vision is still preserved, which makes screening uptake, rather than retinopathy prevalence alone, the operationally decisive indicator [9].

Uptake varies widely across sub-Saharan Africa, with estimates ranging from 21.7% in Ghana to 49.0% in South Africa, suggesting context-specific determinants [10,11]. Ethiopian estimates fall within this range: 31.5% of adults attending two referral hospitals in the north-east had used eye care services in the previous year [12], and 39.6% of patients at a district hospital in the north-west reported a regular eye check-up [13]. These figures contrast sharply with 71.9% reported from Saudi Arabia [14] and 70.5% from Switzerland [15], where organized screening programs and near-universal insurance coverage are in place.

Patient knowledge and attitude are the determinants most frequently implicated in this gap [13,16,17], consistent with the knowledge-attitude-practice-outcome framework that links what patients understand about a condition to the preventive behaviors they adopt [18]. Much of the available Ethiopian evidence comes from regional and district facilities in the north of the country. In Addis Ababa, which holds the largest urban concentration of people with diabetes in Ethiopia, a recent study across five public hospitals found that 55.1% of adults with diabetes had undergone retinal screening in the previous year [19], but no study has examined uptake where specialist eye care is co-located with diabetes follow-up. This gap matters for a specific reason. Menelik II Comprehensive Specialized Hospital houses both a high-volume diabetes follow-up clinic and a national referral center for specialist eye care, with retinal screening delivered on site by trained optometrists five days a week. Uptake at this facility therefore approximates a best-case scenario for physical access, and a low figure would implicate demand-side and care-process barriers rather than the absence of services. In Ethiopia, as elsewhere in sub-Saharan Africa, diabetic retinopathy screening is not organized as a program: there is no standardized national screening pathway, and patients with diabetes are typically linked to eye care only after complications or visual symptoms develop [6,20]. Weak referral links between diabetes clinics and eye services, together with out-of-pocket costs and the lack of recall systems, mean that co-located services do not by themselves guarantee uptake [20]. We conducted this study to determine the prevalence of adequate diabetic retinopathy screening practice, and to identify the factors associated with it, among adults with diabetes on follow-up at this hospital.

## Materials and methods

### Study design, setting and period

An institution-based cross-sectional study was conducted from December 2023 to February 2024 at Menelik II Comprehensive Specialized Hospital, Addis Ababa, Ethiopia. Established in 1903 and administered by the Addis Ababa Health Bureau, the hospital is a national referral center for specialist eye care, staffed by 22 specialists and six subspecialists. Adults with diabetes are followed at the hospital’s Medical Referral Clinic, which operates Monday to Friday with appointment intervals of one week to three months. Diabetes care is delivered by general practitioners, internists and nurses; diabetes health education is provided principally by nurses; and trained optometrists perform diabetic retinopathy screening at the outpatient triage area on all working days. Health Management Information System records showed 4,300 diabetes follow-up visits between January and June 2023, a mean of 717 patients per month. The study is reported in accordance with the STROBE statement for cross-sectional studies (S1 Checklist).

### Participants and eligibility

The source population comprised all adults with diabetes mellitus on follow-up at the Medical Referral Clinic. Patients aged 18 years and above with either type 1 or type 2 diabetes who had been on follow-up for three months or longer were eligible. Patients who were acutely unwell or presenting as emergencies, and women with gestational diabetes, were excluded. Participant flow is summarized in Fig 1.

**Fig 1.**
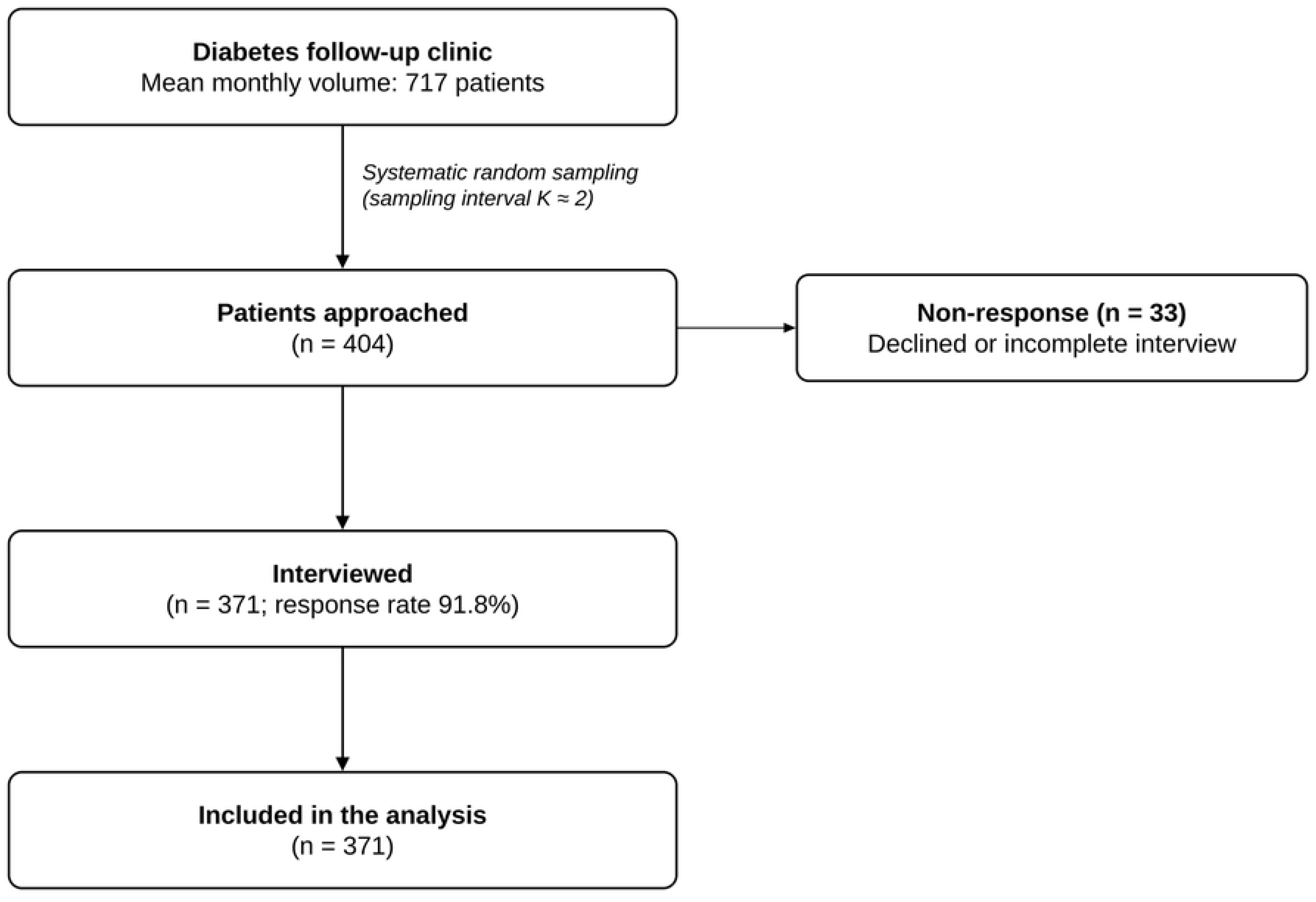
Flow of participants through screening, enrollment and analysis.

### Sample size

The sample size for the primary descriptive objective was calculated using the single population proportion formula, n = Z²₁₋α/₂ p(1 − p)/d², with a 95% confidence level (Z = 1.96), an absolute precision (d) of 5%, and p = 0.396, the proportion of patients reporting a regular eye check-up in the most closely comparable Ethiopian study, conducted at Debark Hospital [13]. This yielded 367 participants; allowing 10% for non-response gave a required sample of 404.

Sample sizes were also computed for the analytical objective using the double population proportion formula in Epi Info StatCalc version 7.2.5.0 (Centers for Disease Control and Prevention, Atlanta, GA, USA), at 95% confidence and 80% power, taking the exposure proportions and odds ratios reported at Debark Hospital for attitude (23.0% exposed among controls, odds ratio 7.46), previous eye disease (20.9%, odds ratio 7.07) and knowledge of diabetic retinopathy (24.5%, odds ratio 6.79) [13]. These gave requirements of 48, 51 and 51 participants respectively, all substantially smaller than the descriptive estimate. The largest figure, 404, was therefore adopted as the final sample size, which conferred more than 80% power for each of the hypothesized associations.

### Sampling procedure

Participants were selected by systematic random sampling in the order of their clinic appointments. Using the mean monthly clinic volume of 717 patients as the denominator and the required sample of 404, the sampling interval was calculated as K = 717/404 ≈ 2. Every second eligible patient exiting the outpatient department was therefore approached, and recruitment continued on consecutive working days until the required sample size was reached.

### Variables and measurement

The outcome variable was diabetic retinopathy screening practice. It was measured with three items adapted from a validated knowledge, attitude and practice instrument [16]: whether the respondent had a regular (at least annual) eye check-up; how often they attended for a dilated eye examination; and how long after the diagnosis of diabetes their first dilated eye examination took place. Each item was scored as correct (1) or incorrect (0) against the recommended standard, giving a total score of 0–3. Participants meeting at least two of the three items were classified as having adequate screening practice, and the remainder as having inadequate practice. The three practice items showed high internal consistency (Cronbach’s α = 0.91).

Knowledge was assessed with 10 items adapted from the Debark Hospital study [13], covering the ocular effects of diabetes, the nature of diabetic retinopathy, its risk factors, and the recommended timing of screening. Each knowledge item was coded as correct (1) or incorrect/don’t know (0), giving a total score of 0–10. Attitude was assessed with six five-point Likert items (1 = strongly disagree to 5 = strongly agree) adapted from the instrument of Srinivasan et al. [16], giving a total score of 6–30. Both scales were dichotomized at the same 80% cut-off: adequate versus inadequate knowledge (≥ 8 of 10) and favorable versus unfavorable attitude (total score ≥ 24 of 30). Internal consistency in this sample was good for the knowledge scale (Cronbach’s α = 0.85) and very high for the attitude scale (α = 0.98), the latter reflecting limited differentiation between attitude items. Independent variables were selected a priori on the basis of the knowledge-attitude-practice-outcome framework [18] and prior Ethiopian studies [12,13]. They comprised socio-demographic characteristics (age, sex, marital status, educational level, residence, employment, health insurance coverage and usual mode of transport to the hospital) and clinical and behavioral characteristics (type of diabetes, duration since diagnosis, family history of diabetes, comorbidities, known eye disorders, exposure to diabetes education and its source), together with the knowledge and attitude scores.

### Data collection and quality control

The questionnaire was developed in English, translated into Amharic and back-translated to check consistency of meaning. It was pre-tested one week before data collection on 10% of the calculated sample size at Zewditu Memorial Hospital, and ambiguous items were revised before fieldwork began. Three nurses holding a Bachelor of Science degree and with at least one year of clinical experience at the Medical Referral Clinic collected the data. They received a one-day training session covering the study objectives, the questionnaire and interview technique. Interviews were conducted face to face as patients exited the outpatient department, and responses were entered directly into the KoBo Toolbox platform, which applied range and skip-pattern checks at the point of entry. The principal investigator supervised data collection daily and reviewed completed questionnaires for consistency.

### Statistical analysis

Data were exported to IBM SPSS Statistics version 26 (IBM Corp., Armonk, NY, USA) for cleaning and analysis. Categorical variables are summarized as frequencies and percentages, and continuous variables as medians with interquartile ranges, since their distributions were skewed. The prevalence of adequate screening practice is presented with a 95% Wilson score confidence interval.

Binary logistic regression was used to examine the association between each independent variable and adequate screening practice. Variables associated with the outcome at p < 0.25 in bivariable analysis were entered simultaneously into a multivariable model; attitude was retained a priori because of its centrality to the conceptual framework, despite not meeting this screening threshold.

Multicollinearity was assessed using the variance inflation factor, with all values below 5 and therefore acceptable. Model fit was assessed with the Hosmer-Lemeshow goodness-of-fit test; a p-value above 0.05 was taken to indicate adequate fit. Discrimination was assessed with the area under the receiver operating characteristic curve. Crude and adjusted odds ratios are reported with 95% confidence intervals, and statistical significance was declared at a two-sided p-value below 0.05.

### Ethics statement

Ethical approval was obtained from the Institutional Review Board of the Addis Continental Institute of Public Health (ACIPH) (Ref. No. ACIPH-MPH/010/16; December 8, 2023) and the Addis Ababa Public Health and Emergency Management Directorate (Ref. No. AAH/9013/227; December 22, 2023). Written informed consent was obtained from every participant after the purpose, procedures, risks and benefits of the study had been explained. Participation was voluntary, and participants were informed of their right to decline, to skip any question and to withdraw at any point without any effect on their care. Medical record numbers were used to link questionnaires during data collection, and no personal identifiers were retained in the analytical dataset. Access to the data was restricted to the investigators, and the study was conducted in accordance with the Declaration of Helsinki.

## Results

### Participant characteristics

Of the 404 patients approached, 371 completed the interview, giving a response rate of 91.8%; the remaining 33 declined or did not complete the interview. All 371 participants were included in the analysis (Fig 1). The median age was 55 years (interquartile range 43–66) and 187 participants (50.4%) were men. Most lived in Addis Ababa (308, 83.0%) and held health insurance (263, 70.9%), and almost half had attended higher education (183, 49.3%). Type 2 diabetes accounted for 289 participants (77.9%), and the median duration since diagnosis was 9 years (interquartile range 6–14). Comorbidity was common, reported by 204 participants (55.0%), of whom 129 (63.2%) had hypertension. A known eye disorder was reported by 151 participants (40.7%), most often cataract (62 of 151, 41.1%). Almost all participants (357, 96.2%) had received information about diabetes and its complications, most commonly from a health professional at the diabetes clinic (254 of 357, 71.1%), and 336 (90.6%) were aware that diabetes can affect the eye. Full participant characteristics are shown in Table 1.

**Table 1.** Socio-demographic, clinical and behavioral characteristics of adults with diabetes mellitus attending Menelik II Comprehensive Specialized Hospital, Addis Ababa, Ethiopia, 2023–2024 (n = 371).

| Characteristic | Frequency (n) | Percentage (%) |
|---|---|---|
| **Socio-demographic characteristics** |  |  |
| Age, years, median (IQR) | 55 (43–66) |  |
| *Age group* |  |  |
| 18–40 | 82 | 22.1 |
| 41–65 | 190 | 51.2 |
| ≥ 66 | 99 | 26.7 |
| *Sex* |  |  |
| Male | 187 | 50.4 |
| Female | 184 | 49.6 |
| *Marital status* |  |  |
| Currently married | 165 | 44.5 |
| Not currently married | 206 | 55.5 |
| *Educational level* |  |  |
| No formal education | 55 | 14.8 |
| Primary | 27 | 7.3 |
| Secondary or vocational | 106 | 28.6 |
| Higher education | 183 | 49.3 |
| *Residence* |  |  |
| Addis Ababa | 308 | 83.0 |
| Outside Addis Ababa | 63 | 17.0 |
| *Employment status* |  |  |
| Employed | 206 | 55.5 |
| Not employed or retired | 165 | 44.5 |
| *Health insurance coverage* |  |  |
| Yes | 263 | 70.9 |
| No | 108 | 29.1 |
| *Usual mode of transport to hospital<sup>a</sup>* |  |  |
| Bus or taxi | 272 | 73.3 |
| Walking | 163 | 43.9 |
| Accompanied by another person | 31 | 8.4 |
| Own vehicle | 18 | 4.9 |
| **Clinical and behavioral characteristics** |  |  |
| *Type of diabetes* |  |  |
| Type 1 | 82 | 22.1 |
| Type 2 | 289 | 77.9 |
| Duration of diabetes, years, median (IQR) | 9 (6–14) |  |
| *Family history of diabetes* |  |  |
| Yes | 143 | 38.5 |
| No | 142 | 38.3 |
| Not known | 86 | 23.2 |
| *Any comorbidity* |  |  |
| Yes | 204 | 55.0 |
| No | 167 | 45.0 |
| *Comorbidity type (n = 204)<sup>a</sup>* |  |  |
| Hypertension | 129 | 63.2 |
| Cardiovascular disease | 37 | 18.1 |
| Respiratory disease | 34 | 16.7 |
| Other | 61 | 29.9 |
| *Known eye disorder* |  |  |
| Yes | 151 | 40.7 |
| No | 220 | 59.3 |
| *Type of eye disorder (n = 151)<sup>a</sup>* |  |  |
| Cataract | 62 | 41.1 |
| Glaucoma | 45 | 29.8 |
| Reduced vision | 31 | 20.5 |
| Diabetic retinopathy | 24 | 15.9 |
| *Received information on diabetes complications* |  |  |
| Yes | 357 | 96.2 |
| No | 14 | 3.8 |
| *Main source of that information (n = 357)* |  |  |
| Health professional at the diabetes clinic | 254 | 71.1 |
| Television, magazines or other media | 41 | 11.5 |
| Family member, relative or friend with diabetes | 29 | 8.1 |
| Ophthalmologist or optometrist | 33 | 9.2 |
| Aware that diabetes can affect the eye | 336 | 90.6 |

IQR, interquartile range. <sup>a</sup> Multiple responses were permitted, so percentages sum to more than 100%. Percentages for subgroups are calculated using the subgroup denominator indicated in the row heading; all other percentages use n = 371.

### Knowledge of and attitude towards diabetic retinopathy screening

Adequate knowledge of diabetic retinopathy and its screening was recorded in 230 of 371 participants (62.0%, 95% confidence interval 57.0–66.8), and a favorable attitude towards screening in 270 (72.8%, 95% confidence interval 68.0–77.1).

### Diabetic retinopathy screening practice and reported barriers

Adequate diabetic retinopathy screening practice was recorded in 139 of 371 participants (37.5%, 95% confidence interval 32.7–42.5); the remaining 232 (62.5%) had inadequate practice. Of the 139 participants with adequate practice, 132 (95.0%) met all three practice items. A larger group, 210 participants (56.6%), reported having a regular (annual) eye check-up; of these, 108 (51.4%) were examined by an ophthalmologist at the eye hospital and 102 (48.6%) by a physician.

Of the 161 participants without a regular eye check-up, 160 answered the question on barriers to attending eye check-ups. The cost of the examination was cited most frequently, by 72 (45.0%), followed by lack of time (47, 29.4%), long travel distance (43, 26.9%), lack of transport (29, 18.1%) and forgetting the appointment (14, 8.8%); 31 (19.4%) cited other reasons (Fig 2).

**Fig 2.**
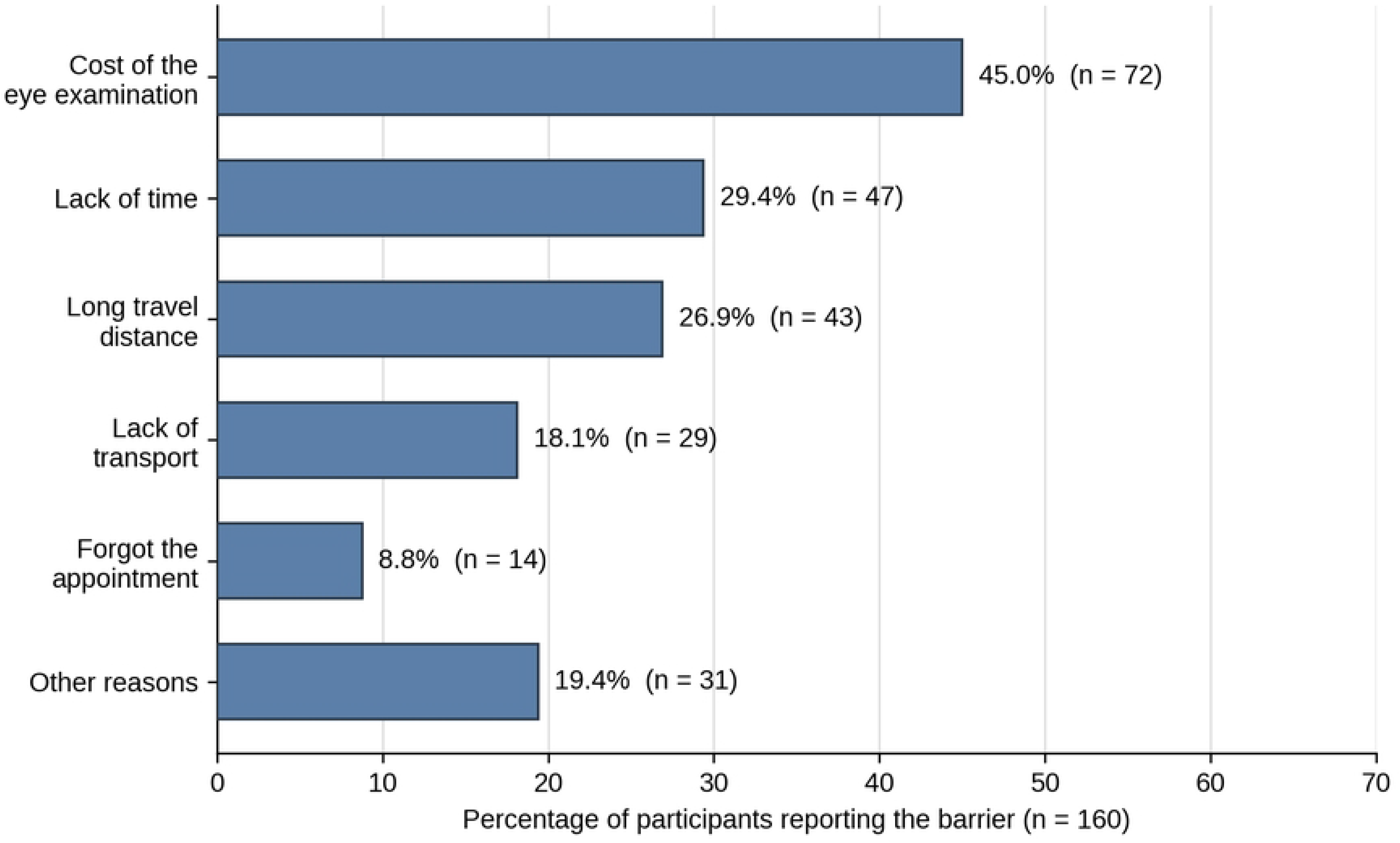
Barriers to attending eye check-ups among participants without a regular eye check-up (n = 160). Note: *Multiple responses were permitted, so percentages sum to more than 100%*.

### Factors associated with diabetic retinopathy screening practice

In bivariable analysis, age, marital status, educational level, employment status, health insurance coverage, type of diabetes, a known eye disorder and knowledge of diabetic retinopathy screening were associated with adequate screening practice at p < 0.25 and were entered into the multivariable model, together with attitude, which was retained a priori.

After adjustment, five variables remained independently associated with adequate screening practice. Compared with participants aged 18–40 years, those aged 41–65 years had more than three times the odds of adequate practice (adjusted odds ratio 3.15, 95% confidence interval 1.10– 9.03, p = 0.033), whereas the odds among those aged 66 years and above did not differ significantly from the youngest group. Currently married participants had higher odds than those who were not currently married (1.73, 1.01–2.97, p = 0.044), and participants who had attended higher education had almost four times the odds of those with no formal education (3.82, 1.33–10.95, p = 0.013). A known eye disorder was associated with nearly a fourfold increase in the odds of adequate practice (3.96, 2.11–7.43, p < 0.001), and adequate knowledge with a fivefold increase (5.10, 2.81–9.25, p < 0.001). A favorable attitude was inversely associated with adequate practice (0.36, 0.19–0.68, p = 0.002), an association that was not evident before adjustment (crude odds ratio 0.94, 0.58–1.50). The model showed adequate fit (Hosmer-Lemeshow χ² = 7.23, df = 8, p = 0.51) and acceptable discrimination (area under the curve 0.79). Full results are presented in Table 2 and S1 Table.

**Table 2.** Bivariable and multivariable logistic regression of factors associated with adequate diabetic retinopathy screening practice among adults with diabetes mellitus, Addis Ababa, Ethiopia, 2023–2024 (n = 371).

| **Characteristic** | **Adequate practice, n (%)** | **Crude OR (95% CI)** | **Adjusted OR (95% CI)** | **p-value** |
|---|---|---|---|---|
| *Age group, years* |  |  |  |  |
| 18–40 | 22 (26.8) | 1 (reference) | 1 (reference) |  |
| 41–65 | 91 (47.9) | 2.51 (1.42–4.41) | 3.15 (1.10–9.03) | 0.033* |
| ≥ 66 | 26 (26.3) | 0.97 (0.50–1.88) | 2.66 (0.77–9.23) | 0.122 |
| *Marital status* |  |  |  |  |
| Not currently married | 60 (29.1) | 1 (reference) | 1 (reference) |  |
| Currently married | 79 (47.9) | 2.24 (1.46–3.43) | 1.73 (1.01–2.97) | 0.044* |
| *Educational level* |  |  |  |  |
| No formal education | 8 (14.5) | 1 (reference) | 1 (reference) |  |
| Primary | 8 (29.6) | 2.47 (0.81–7.55) | 1.50 (0.42–5.42) | 0.536 |
| Secondary or vocational | 38 (35.8) | 3.28 (1.41–7.67) | 2.47 (0.89–6.86) | 0.082 |
| Higher education | 85 (46.4) | 5.10 (2.28–11.39) | 3.82 (1.33–10.95) | 0.013* |
| *Employment status* |  |  |  |  |
| Not employed or retired | 45 (27.3) | 1 (reference) | 1 (reference) |  |
| Employed | 94 (45.6) | 2.24 (1.44–3.47) | 1.48 (0.76–2.88) | 0.255 |
| *Health insurance coverage* |  |  |  |  |
| Yes | 88 (33.5) | 1 (reference) | 1 (reference) |  |
| No | 51 (47.2) | 1.78 (1.13–2.81) | 1.58 (0.86–2.89) | 0.141 |
| *Type of diabetes* |  |  |  |  |
| Type 1 | 26 (31.7) | 1 (reference) | 1 (reference) |  |
| Type 2 | 113 (39.1) | 1.38 (0.82–2.33) | 0.54 (0.19–1.55) | 0.252 |
| *Known eye disorder* |  |  |  |  |
| No | 70 (31.8) | 1 (reference) | 1 (reference) |  |
| Yes | 69 (45.7) | 1.80 (1.18–2.77) | 3.96 (2.11–7.43) | < 0.001* |
| *Knowledge of diabetic retinopathy screening* |  |  |  |  |
| Inadequate | 25 (17.7) | 1 (reference) | 1 (reference) |  |
| Adequate | 114 (49.6) | 4.56 (2.76–7.55) | 5.10 (2.81–9.25) | < 0.001* |
| *Attitude towards diabetic retinopathy screening* |  |  |  |  |
| Unfavorable | 39 (38.6) | 1 (reference) | 1 (reference) |  |
| Favorable | 100 (37.0) | 0.94 (0.58–1.50) | 0.36 (0.19–0.68) | 0.002* |

*CI, confidence interval; OR, odds ratio. Percentages are row percentages, calculated within each category. Adjusted odds ratios are from a single multivariable logistic regression model containing all variables shown. Model fit was acceptable (Hosmer-Lemeshow χ² = 7.23, p = 0.51; area under the curve 0.79) and no variance inflation factor exceeded 5. \* Statistically significant at p < 0.05.*

## Discussion

In this study, just over one-third of participants had undergone retinal examination at the recommended frequency. This shortfall is striking given that diabetes follow-up and a national referral center for specialist eye care operate within the same compound, with retinal screening available five days a week. Adequate knowledge of diabetic eye disease was the strongest correlate of screening uptake, followed by the presence of a known eye disorder, higher education, middle age, and being currently married.

The prevalence we observed is similar to the 39.6% reported at Debark Hospital in north-west Ethiopia [13] and to findings from tertiary hospitals in India [16] and Bangladesh [17]. It is somewhat higher than the 31.5% found in two referral hospitals in north-east Ethiopia [12], yet substantially lower than the 71.9% and 70.5% reported from Saudi Arabia [14] and Switzerland [15]. The latter settings differ in ways that plausibly explain the gap: both operate organized recall systems with defined screening intervals, and both have near-universal insurance coverage that removes the point-of-service cost. The more informative comparison, however, is local. Uptake at this hospital, where specialist eye care is available on site, was no higher than at Debark Hospital and was lower than the 55.1% reported across five other public hospitals in Addis Ababa [19], although that study measured a single dilated examination in the previous year rather than a composite of recommended practices. This suggests that the availability of screening capacity is not, by itself, the binding constraint. Attention should instead fall on how patients are identified, counseled and referred within the diabetes clinic.

Participants aged 41–65 years had more than three times the odds of adequate screening practice compared with those aged 18–40 years, consistent with findings from north-east Ethiopia [12], Hawassa [21] and South Africa [11]. Age-related ocular morbidity accumulates during this period, leading to more frequent contact with eye services [22]. In contrast, younger adults may have less flexibility to attend appointments because of work and study commitments [23]. The lack of a significant association in the oldest age group is notable and may reflect a countervailing effect: reduced mobility and greater dependence on others for transport can offset the higher clinical need of older patients. This interpretation aligns with the prominence of transport-related barriers in our data.

Currently married participants had higher odds of adequate screening practice than those who were not, consistent with evidence that spousal support improves uptake of preventive services in general [24]. Educational attainment showed a clear graded relationship: participants with higher education had almost four times the odds of adequate practice compared with those who had no formal schooling. This pattern has also been observed at Felege Hiwot Specialized Hospital in Ethiopia [25] and in the Swiss CoDiab-VD cohort [15]. Higher health literacy is likely to improve both the comprehension of screening advice and the capacity to navigate referral pathways.

The strong association with a known eye disorder carries the clearest public-health message. Participants who already had an ocular diagnosis had almost four times the odds of adequate screening practice. This suggests that eye examination in this population is often prompted by existing ocular problems rather than undertaken as routine prevention, although the cross-sectional design cannot exclude the reverse explanation that eye disorders were detected because participants attended. Similar patterns have been described in north-east Ethiopia [12], at Debark Hospital [13] and in southern India [26]. This is precisely the pattern that screening programs are designed to reverse, because the window in which treatment can preserve vision closes before symptoms appear.

Adequate knowledge was associated with a fivefold increase in the odds of adequate practice, consistent with studies from Kenya [27], Turkey [28], Bangladesh [17], Indonesia [29] and the United Kingdom [30]. Because knowledge is modifiable and because almost all participants (96.2%) had already received information on diabetes and its complications, most often from clinic staff, this represents the most tractable point of intervention: the content of existing counseling, rather than its reach, appears to be the limiting factor.

The inverse association between a favorable attitude and adequate screening practice is counter-intuitive and warrants cautious interpretation. The relationship was absent in the unadjusted analysis and only emerged after multivariable adjustment, a pattern that typically signals residual confounding or collinearity, most plausibly with knowledge, given the strong inter-correlation of these constructs in knowledge–attitude–practice scales. Compounding this, the attitude measure suffered from a pronounced ceiling effect, with nearly three-quarters of participants classified as favorable, thereby reducing its ability to differentiate behavior. Beyond these methodological limitations, a substantive explanation remains plausible: even participants who value screening may be unable to act on that preference when confronted with the practical barriers of cost, time, and distance that participants most often reported. Similar gaps between positive attitudes and actual attendance have been reported in Saudi Arabia [31] and in northern India [32]. We therefore view this finding as hypothesis-generating, underscoring the need for future work that disentangles attitudinal measures from the structural constraints that ultimately shape screening behavior.

These results point to three practical priorities. First, diabetic retinopathy screening should be offered opportunistically at every diabetes follow-up visit, using a documented annual due-date in the clinic record. Second, given that the optometry service operates within the same hospital, a direct internal referral pathway from the medical referral clinic to the triage screening point would eliminate the need for a separate patient journey. Third, counseling should be restructured to address the specific knowledge gaps identified here, particularly the asymptomatic nature of treatable retinopathy. Cost was the barrier most often reported, yet uninsured participants were more likely than insured participants to have adequate screening practice (47.2% vs 33.5%), a difference that was not statistically significant after adjustment. This may reflect limited coverage of eye examinations under current insurance schemes or differences in the characteristics of uninsured patients attending this hospital, and the role of insurance benefit design in screening uptake warrants further study.

### Strengths and limitations

The strengths of this study include a sample size adequate for the planned analyses, a high response rate, systematic random selection of participants, use of an instrument adapted from previously published and locally pre-tested tools, and the fact that it provides the first estimate of screening practice from an Ethiopian facility in which specialist eye care is co-located with diabetes follow-up.

Several limitations should be considered. The cross-sectional design precludes any inference about the direction of the observed associations, particularly for knowledge and attitude, which may equally be consequences of prior screening attendance. Screening practice was self-reported, and interviews were conducted by facility nurses, so recall and social desirability bias are likely. The study was conducted at a single tertiary facility with co-located specialist eye services and a predominantly urban, relatively well-educated population, so the findings are unlikely to generalize to rural or primary care settings. Clinical measures known to influence both retinopathy risk and care-seeking, notably glycemic control, were not available, so residual confounding cannot be excluded. Age and diabetes type were closely related (70 of the 82 participants with type 1 diabetes were aged 18–40 years), so the adjusted estimates for these two variables should be interpreted with caution. Responses to the attitude items were highly uniform, with most participants giving the same response to all six items; this limited the scale’s ability to distinguish between participants and may partly explain the unexpected direction of the attitude association.

## Conclusions

Fewer than four in ten adults with diabetes attending a tertiary hospital in Addis Ababa underwent diabetic retinopathy screening at the recommended frequency, despite the availability of on-site specialist eye care. Screening was strongly associated with having a known eye disorder, suggesting that eye examination is often prompted by existing ocular problems rather than undertaken for prevention. Adequate knowledge of diabetic retinopathy was the strongest modifiable factor associated with screening uptake. Integrating diabetic retinopathy screening into routine diabetes follow-up, establishing a streamlined referral pathway to on-site eye services, and strengthening patient counseling to emphasize the asymptomatic nature of early disease may improve screening uptake more effectively than general awareness campaigns alone. Future studies should verify screening attendance using clinical records and include primary care settings and health-care provider perspectives to better inform strategies for improving screening coverage.

## Data Availability

The data underlying the findings reported in this study are publicly available in Figshare at: https://figshare.com/s/33c957e8893940dc9745?file=71092764

https://figshare.com/s/33c957e8893940dc9745

## Acknowledgments

We thank the staff of the Medical Referral Clinic at Menelik II Comprehensive Specialized Hospital for facilitating data collection, the data collectors for their careful work, and all the study participants for the time they gave to this research.

## Supporting information

**S1 Checklist. STROBE checklist for cross-sectional studies.**

**S1 Table. Full multivariable logistic regression model for adequate diabetic retinopathy screening practice.** Regression coefficients, standard errors, Wald statistics, p-values, adjusted odds ratios with 95% confidence intervals, and model fit statistics (n = 371).

